# Eye-to-Eye: Gaze detection as a proxy for medical doctor behaviour during virtual and in-person appointments

**DOI:** 10.64898/2026.11.17.26368012

**Authors:** Ricardo Antaõ, Plinio Moreno, José Santos-Victor, Nuno André da Silva, Raquel Santos, Francisca Leite, Daniel Ferreira

## Abstract

The gaze direction of the physician during consultations plays an important role in patient satisfaction, with several studies pointing to the positive correlation between the amount of time the physician spends looking at the patient and patient satisfaction. Additionally, the effects of the virtual consultation environment on the gaze behaviour of the doctor have yet to be studied. Therefore, this study aims to assess the impact of the virtual consultation environment by comparing the amount of total time the doctor spends looking at the patient between face-to-face and virtual consultations, as well as the average duration during the consultation and the gaze frequency. The study population consisted of 14 doctors divided between 4 medical specialties: Gynaecology/Obstetrics (4), Neurology (3), Endocrinology (3) and General and Familiar Medicine (4). A low-cost desktop that runs an appearance-based gaze estimation pipeline was implemented in the clinical setup. Each doctor recorded 20 face-to-face consultations and 20 virtual consultations, which were then processed by the pipeline to obtain the percentage of time the doctor spent looking at the patient during the consultation. Overall, we found that doctors look more at the patient in virtual consultations compared to face-to-face consultations, both in total time and average duration during the consultation. Within medical specialties, three out the four specialties involved presented clear tendencies to look more at the patient in virtual consultations when compared to face-to-face consultations.

## I. Introduction

The physician-patient relationship is a crucial component of the effectiveness of any health care system. Effective communication, encompassing both verbal and non-verbal cues, stands as a cornerstone of a strong physician-patient relationship. Within this dynamic, the dialogue exchanged during medical interviews assumes a paramount role [1]–[5].

Communication in the physician-patient relationship is divided into verbal and non-verbal communication. Verbal communication is defined as communication behaviour with linguistic content [3]. This is classified according to the model described by Bird and Cohen-Cole model [6]. According to this model, we can classify verbal interactions into three key functions: data gathering to understand the patient (gathering information), development of a rapport and responding to the patient’s emotions (developing therapeutic relationship), and patient education and behavioural management (decision making and management). Non-Verbal communication can be defined as communication behaviour without linguistic content and is typically distinguished by which part of the body is being used to express the behaviour. Face non-verbal behaviours include smiling, gazing, frowning, eyebrow-raising, and facial expressivity. Body non-verbal behaviour is expressed through posture or gestures. Vocal non-verbal behaviour includes loudness, voice pitch, monotony, and speech rate. Several studies indicate that the non-verbal cues of the physician are one of the most critical aspects of physician-patient communication [2]–[5].

Among the non-verbal behaviour, the gaze is one of the most important cues to analyze in non-verbal communication. Studies have concluded that there is a positive correlation between patient satisfaction and the amount of eye contact between the physician and the patient [5]. Larger eye contact time leads to increased warmth and competence in the patient-doctor interaction [7]. The amount of time the physician is gazing at the patient and not at the patient’s health records on the screen can improve the patient’s perception and cognitive functioning [8]. Additionally, the rise in the use of virtual consultations due to the COVID-19 pandemic provided an entirely different environment for the physician-patient relationship. Its impact in the physician-patient relationship is yet to be fully understood.

Moreover, studying and quantifying non-verbal communication is also challenging. The majority of the studies on this topic use manual annotation systems to quantify non-verbal behaviours. This process is costly and laborious, which is not convenient or scalable. Recently, some methods to automate annotations have been proposed [8], [9]. However, they were designed for a very constrained environment which would not translate well to other consultation offices/setups. Nonetheless, they provide the first approaches to automatic classification of physician’s gaze during medical appointments.

In the current study, we focus on the automated analysis of the gaze direction of the physician during consultations, classifying it according to whether the physician is looking at the patient or not. It will aim to implement a scalable and low-cost appearance-based gaze estimation system in the clinical setup to support the analysis of the physician-patient relationship. In order to demonstrate the methodology and its potential, a study to quantify the face-to-face vs virtual consultations was conducted.

## II. Related Work

### A. Gaze Estimation for images

Gaze estimation aims to compute the subject’s gaze direction from images. The earliest attempts at gaze estimation consisted in the detection of eye movement patterns like fixation, saccades and smooth pursuits [10]. These early methods attached movement sensors around the eyes to detect the movement patterns mentioned before, which were invasive and not scalable due to specific hardware requirements. However the evolution of computer vision technology enabled the creation of modern eye tracking software devices. Of the various modern gaze estimation methods developed in recent years, deep learning powered appearance-based gaze estimation systems are the methods that provide state-of-the-art accuracy in a robust manner (apart from proprietary commercial eye trackers).

Appearance-based gaze estimation methods learn functions that map directly from images to gaze direction by using image features like image pixels [11] or deep features [12] and are able to work with off-the-shelf web cameras. Regressing gaze directly from images is a difficult task due to the variety and complexity of eye appearance and several models were tested. However over recent years, deep learning methods have been found to be the most effective for the task with the majority of gaze estimation research today focusing on deep learning approaches [12]–[22].

*Gaze360*, is one of the state-of-the-art methods developed with deep learning techniques [15]. It aimed to provide robust gaze estimation in unconstrained environments with a wide variety of head poses and illumination conditions. To achieve this, they propose both a dataset and a deep learning gaze estimation model, both denominated of *Gaze360*. The Gaze360 model is a video-based gaze tracking model using a bidirectional Long-Short Term Memory (LSTM) [23], in conjunction with a backbone network and a fully connected layer. The model achieves very robust results, being able to provide state-of-the-art accuracy under unconstrained conditions like, extreme head poses or illumination changes.

### B. Gaze Estimation on a clinical setup

The authors in [8], proposed a gaze classification approach for doctor’s gaze using convolutional neural networks (CNNs). They used the VGG-16 architecture [24] pre-trained on the ImageNet dataset, and added to it one Global Max Pooling layer, one Dropout layer and five fully connected layers. They then fine tuned the architecture with a dataset of raw videos from clinical interactions developed by the authors of the study. This dataset consisted of a set of 101 clinical interactions involving 10 doctors and 101 patients. Each interaction was comprised of 3 videos captured at the same time from 3 different cameras, a *Patient-Centered* camera focused on the patient, a *Doctor-Centered* camera focused on the doctor and a *Wide-frame* camera providing wide-view image of both the doctor and patient. The videos were annotated using the Noldus Observer XT Software [25]. In the end, the model proved to be very accurate with a 98.31% accuracy on the validation set and over 80% accuracy on the majority of independent hold out sets with unseen doctors and interactions. However, the need to use 3 different cameras with specific views of the consultation office makes this system complex and unfeasible to scale in many situations. In addition to this, it is unknown how the accuracy of the method translates to different consultation setups with different conditions, *e*.*g*. lighting conditions, patient positioning.

The authors in [26], use eye-tracking glasses to track the gaze of the doctor during face-to-face consultations. A total of 16 doctors seeing a total of 100 patients, each doctor seeing between 2 and 14 patients with the median being 6 patients. Doctor’s gaze was measured with 3 different metrics: *face gaze duration, face gaze frequency* and *face gaze dwell time. Face gaze duration* corresponds to the total amount of time per minute the doctor spent looking at the patient, *face gaze frequency* corresponds to the amount of times per minute the doctor’s gaze switched to the patient and *face gaze dwell time* is the time the doctor’s gaze dwelled on the patients in each instance the doctor looked at them. The study concluded that there was a significant positive correlation between *face gaze dwell time* and *face gaze duration*, a significant negative correlation between *face gaze dwell time* and *face gaze frequency* and no correlation between *face gaze frequency* and *face gaze duration*. Additionally, the study also concluded that the amount of face gaze present in the beginning part of the consultation had positive and significant association to the amount of face gaze present in the beginning of the consultation. It also found that the amount of time the doctor spends looking at the patient during a consultation decreases in the final parts of the consultation compared to the beginning of the consultation.

## III. Experimental Design

With the increase of video consultation there is no study that quantifies and compares video with in-person consultation regarding eye contact between patient and physician in a non-invasive manner. Our study aims to quantify and compare the amount of time the doctor spends looking at the patient between face-to-face and virtual consultations.

### A. Hypotheses

We consider three characteristics to study the differences between in-person and virtual consultations: (i) the percentage of time the doctor spent looking at the patient during a consultation (*Patient%*), (ii) The gaze duration and (iii) the gaze frequency. For gaze duration and frequency, the appointment time is divided into 10 deciles. For each decile, the time spent looking at the patient corresponds to gaze duration, and the number of times the doctor changes the gaze from one of the zones to the patient’s face, corresponds to gaze frequency. We verify three hypotheses between in-person and virtual consultations: (i) the gaze percentage time is similar, (ii) the gaze duration is dependent and (iii) gaze frequency is dependent. We also verify if gaze frequency and gaze duration are dependent for the same type of consultation. These hypotheses are evaluated on the whole doctor population and by specialty.

### B. Study Population

The source population for this study consists of 14 doctors divided among 4 different medical specialties. Choosing doctors among 4 medical specialties increases the scope of our study, at the expense of some robustness in the data. The 4 medical specialties chosen were:

- **Family and General Medicine** - 4 doctors
- **Endocrinology** - 3 doctors
- **Gynecology/Obstetrics** - 4 doctors
- **Neurology** - 3 doctors

The sample size for each doctor was defined as 20 face-to-face and 20 virtual consultations. Doctors from now on will be mentioned in the form of D*X* with *X* being the number assigned to the doctor in the beginning of the study.

### C. Statistical test and Size Effect

To compare the *Patient%* distributions of each doctor the Mann-Whitney U test [27] was used. The *Patient%* distributions were considered to be non-parametric since 20 results are not enough to check the normality of the distributions in most cases. Additionally, since face-to-face and virtual consultations have different participants, *i*.*e*. different patients, the consultation groups were considered to be groups in different conditions and with different participants. A statistical significance was considered for *alpha* of *p <* 0.05.

When a statistically significant difference existed, the Cohen’s *d* size effect [28] was used to calculate the size effect. To verify dependence between gaze frequency and gaze duration between the two types of consultations, we use distance covariance test of independence [29], which finds non-linear dependencies. For instance, to compare the gaze duration for both types of consultations across deciles, the null hypotesis is that the two random variables are independent (*P >* 0.05).

## IV. Experimental set up

### A. Consultation Recording

To record the consultations, an extra camera and computer were setup in the consultation rooms. The conventional consultation room for face-to-face consultations, possesses one computer screen, while the virtual consultation room for virtual consultations, possesses two computer screens. During face-to-face consultations, the patient is present in the room with the doctor while their health records are shown in the computer screen of the conventional consultation room. During virtual consultations, the patient’s video feed is shown on one of the screens of the virtual consultation room while the health records are shown in the other screen.

During both types of consultations, the doctors started and stopped the recordings, corresponding to the beginning and end of each consultation. Due to privacy concerns, only the video feed and no audio was recorded. In this way, only the doctor’s image would be recorded and patient privacy was maintained.

### B. Gaze Estimation Pipeline

The gaze estimation pipeline takes as input a video and the camera’s extrinsic and intrinsic parameters and outputs the frame-by-frame 2D gaze estimates/points of gaze (*PoG*), which are points in the 2D screen plane, illustrated in Figure 2. The pipeline is composed of 3 main blocks illustrated in Figure 1.

**Fig. 1:**
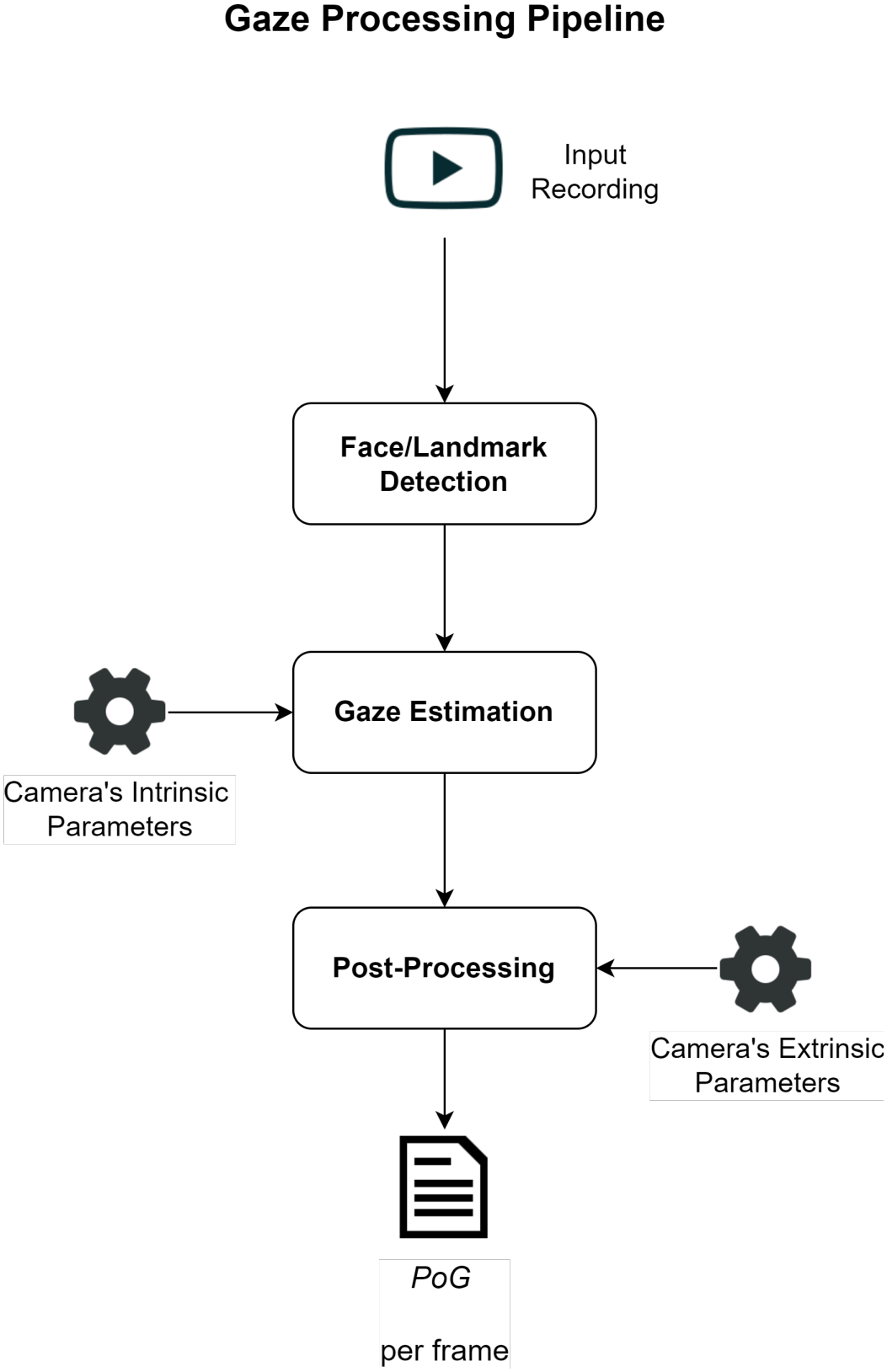
Gaze estimation pipeline composed of 3 main blocks: *Face/Landmark Detection, Gaze Estimation* and *Post-Processing*.

**Fig. 2:**
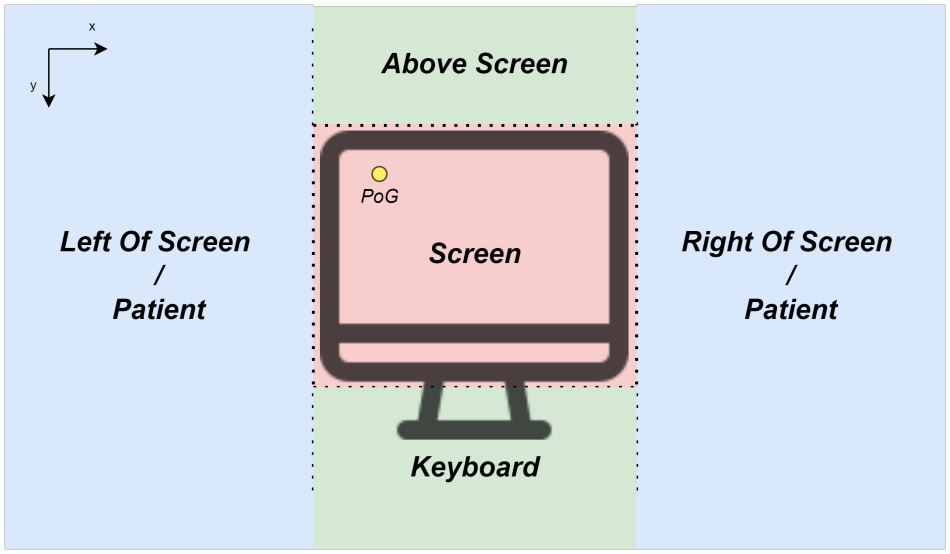
Gaze classification zones: *Above Screen, Screen, Keyboard, Right Of Screen* and *Left Of Screen*. For each consultation room, either *Right Of Screen* or *Left Of Screen* are considered to be the *Patient* zone depending on where the patient is positioned is relation to the screen.

The *Face/Landmark Detection* block serves as the preprocessing of the input video before the gaze estimation step. It takes the input recording and performs the face detection and 2D landmark annotation routines from *3DDFA V2* [30]. The output of the block is the frame-by-frame face bounding box (locating the doctor’s face on the frame) and the 2D landmark annotations of the doctor’s face.

The *Gaze Estimation* block serves to extract the doctor’s gaze direction during the consultation. It takes as input the consultation recording, the camera’s intrinsic parameters and the output of the *Face/Landmark Detection* block. The gaze estimation is performed by the *Gaze360* model [15] and the output of the block is the frame-by-frame 3D gaze estimates/gaze directions.

The *Post-Processing* block converts the 3D gaze estimates/gaze directions into 2D gaze estimates/*PoG*s. It takes as input the 3D gaze estimates from the *Gaze Estimation* block and the camera’s extrinsic parameters. With the camera’s extrinsic parameters, *i*.*e*. the rotation and translation between the Camera Coordinate System (CCS) and the Screen Coordinate System (SCS, the 2D screen plane), we can convert the 3D gaze estimates represented in the CCS to 2D gaze estimates in the SCS. Therefore the output of the block will be the frame-by-frame *PoG*s of the doctor during the consultation, which are going to be classified according to Figure 2.

### C. PoG classification

After obtaining the *PoG*s, we need to classify them in order to distinguish between the doctor looking at the patient and looking at screen/keyboard. Thus, we divided the screen plane into 5 zones as illustrated in Figure 2, *Above Screen, Screen, Keyboard, Right Of Screen* and *Left Of Screen*. In each consultation room the patient could be either on the left or on the right of the screen. Therefore, for each consultation room, either the *Right of Screen* zone or the *Left Of Screen* would be classified as the *Patient* zone. In the end, the classification allowed us to calculate how much time during the consultation the doctor spent looking at the patient by calculating the percentage of *PoG*s in the *Patient* zone. The percentage of *PoG*s classified as *Patient PoG*s was denominated as *Patient%* and it is the variable which is going to be compared in this study.

In face-to-face consultations we assume that if the physician’s face was not visible in the image, the physician is with the patient. This is reasonable because in-person consultation might have physical examination, while it is not the case for virtual consultations.

## V. Results

### A. Global analysis

In total percentage of time, the doctors gaze more at the patient during virtual consultations (55.2%) than in-person consultations (46.5%), (*P <* .001). Regarding gaze duration, Figure 3 shows the average duration of a fixation across all consultation deciles. Note that the trend of gaze duration for the virtual consultations case is decreasing (*β* = *−*0.854, *P <* .001), while the in-person consultations there is no clear trend (*β* = *−*0.11, *P* = 0.262). In-person gaze duration decreases during the first part of the consultation, while increases during the second half of the consultation.

**Fig. 3:**
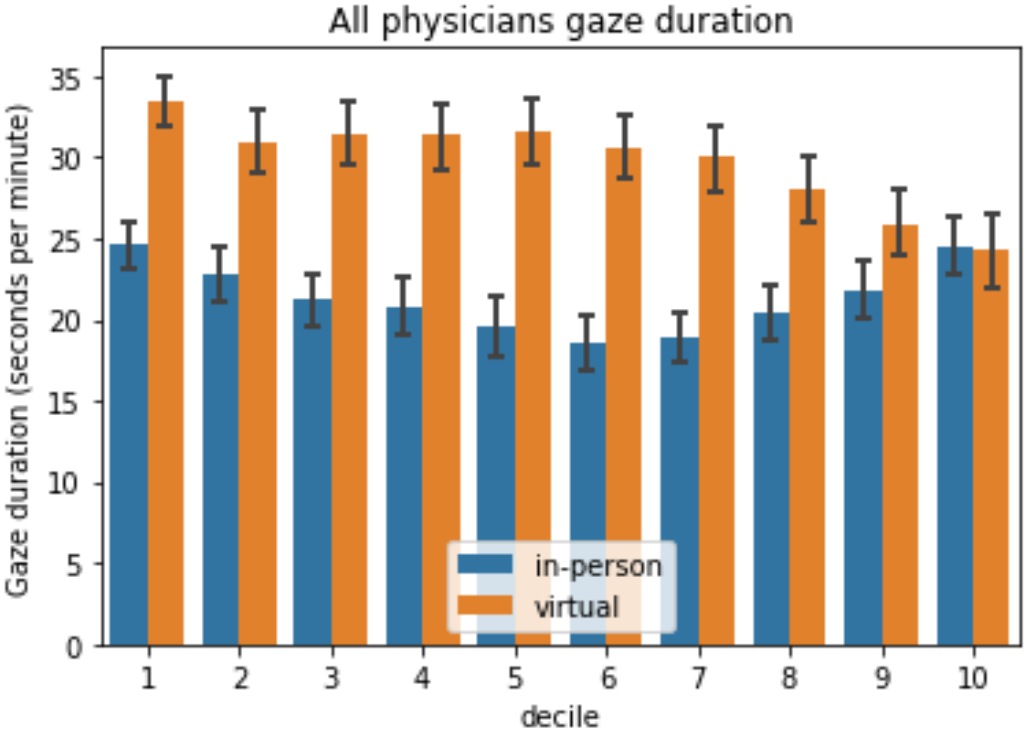
Gaze duration across consultation time. Consultation time is divided into 10 slots.

We wanted to study the gaze duration dependence between both types of consultations. In the case of gaze duration, we found that virtual consultations are independent to in-person ones (*P* = 0.761). We observe that physicians tend to have larger gaze durations during virtual consultations, which happens mostly during the 3-7 deciles. Regarding gaze frequency, Figure 4 shows the average gaze frequency across all consultation deciles. In the case of virtual consultations, the time-series trend is a decreasing one (*β* = *−*0.08, *P* = 0.028). For in-person consultations, the time-series trend of the gaze duration is not clear (*β* = *−*0.0096, *P* = 0.895). In-person gaze duration decreases during the first half of the consultation, while increases during the second half of the consultation.

**Fig. 4:**
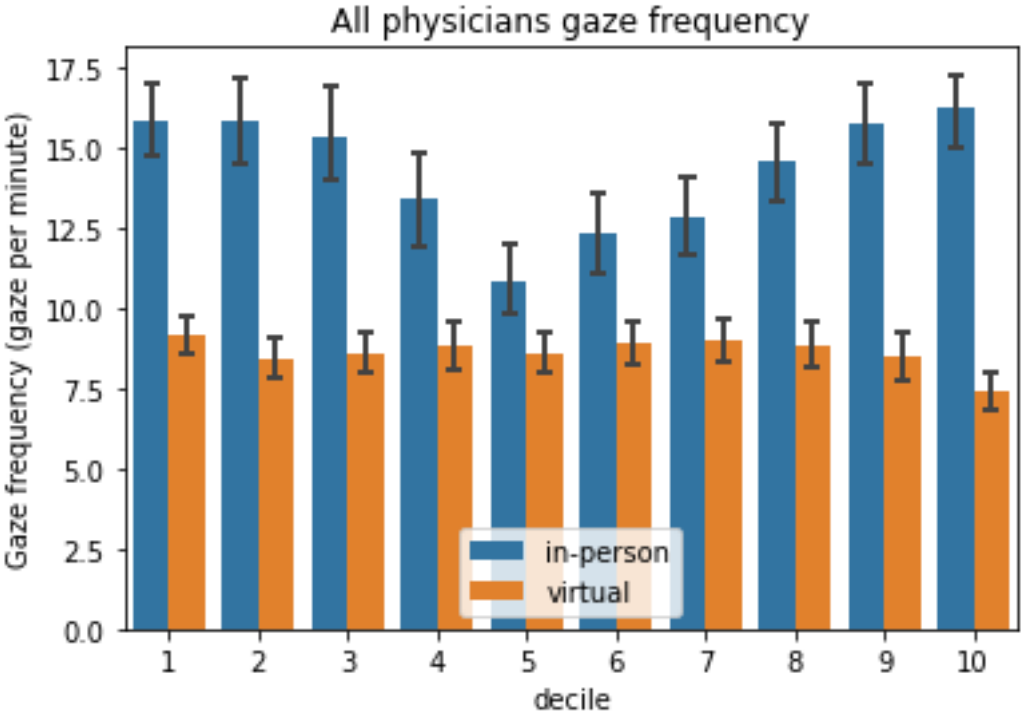
Gaze frequency across consultation time. Consultation time is divided into 10 slots.

We also verify the dependence between gaze frequencies for both conditions. Gaze frequency of virtual consultations is dependent to in-person frequency (*P* = 0.0049). In this case the frequency is almost constant for the virtual consultations.

Finally, we checked if there is a correlation between gaze duration and gaze frequency. For both consultation conditions, there is a significant but relatively small correlation: (i) In-person duration and frequency are correlated (*r* = 0.317, *P <* 0.001), and (ii) virtual duration and frequency are correlated (*r* = 0.118, *P <* 0.001).

### B. Analysys per specialty

#### 1) Gynaecology/Obstetrics

Table I shows the total gaze time to the patient, for each doctor in this specialty and the global values for all doctors. Note that the only significant difference is on D12, while D9 shows a tendency. D6 and D14 do not have differences between the total gaze time. Figure 5 shows the violin plots that illustrate these results. The joint analysis shows a small difference between in-person and virtual gaze times, but most of that difference comes from D12.

**Table I:** Gynaecology/Obstetrics results and summary statistics. *Med*_*f*_ is the median value of total gaze time to the patient for in-person consultations. *Med*_*v*_ is the median of gaze time to the patient for virtual consultations

| Doctor | *Med*<sub>*f*</sub> | *Med*<sub>*v*</sub> | *p-value* | Cohen's *d* |
|---|---|---|---|---|
| D6 | 51.8% | 51.9% | 0.88 | - |
| D9 | 52.3% | 58.9% | 0.06 | - |
| D12 | 55.7% | 75.1% | 1.3 × 10<sup>−5</sup> | 0.69 |
| D14 | 47.9% | 43.4% | 0.56 | - |
| Joint | 51.3% | 57.5% | 0.7 × 10<sup>−2</sup> | - |

**Fig. 5:**
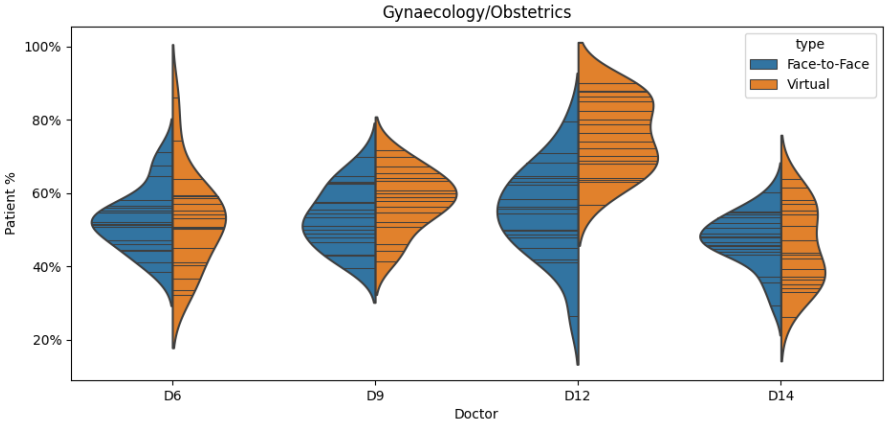
Gynaecology/Obstetrics doctors violin plots

Regarding gaze duration, Figure 6 shows the average duration of a fixation across all consultation deciles. Note that the trend of gaze duration for the virtual consultations case is decreasing (*β* = *−*1.686, *P <* .001), while the in-person consultations there is no clear trend (*β* = *−*0.146, *P* = 0.418). In-person gaze duration decreases during the first part of the consultation, while increases during the second half of the consultation.

**Fig. 6:**
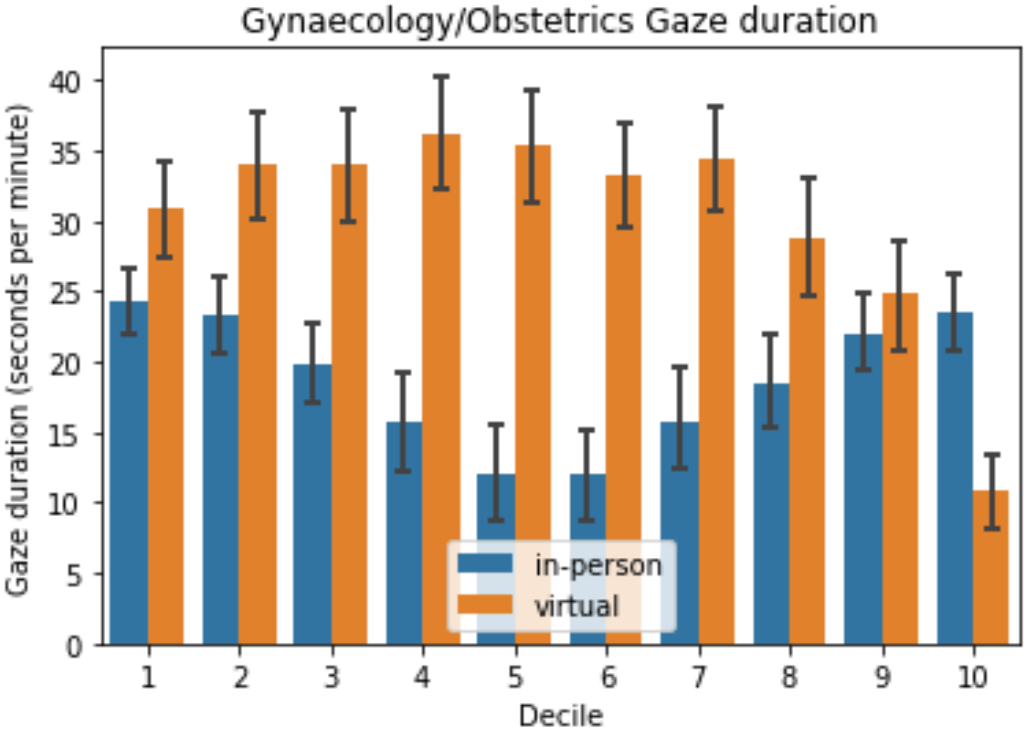
Gaze duration across consultation time for Gynaecology/Obstetrics specialty. Consultation time is divided into 10 slots.

We also verify the dependence between gaze duration for both conditions. Gaze duration of virtual consultations is dependent to in-person frequency (*P* = 0.0298).

Regarding gaze frequency, Figure 7 shows the average gaze frequency across all consultation deciles. In the case of virtual consultations, there is no clear time-series trend (*β* = *−*0.075, *P* = 0.345). Similary, there is no clear trend on in-person consultations (*β* = *−*0.131, *P* = 0.486). In-person gaze duration decreases during the first half of the consultation, while increases during the second half of the consultation.

**Fig. 7:**
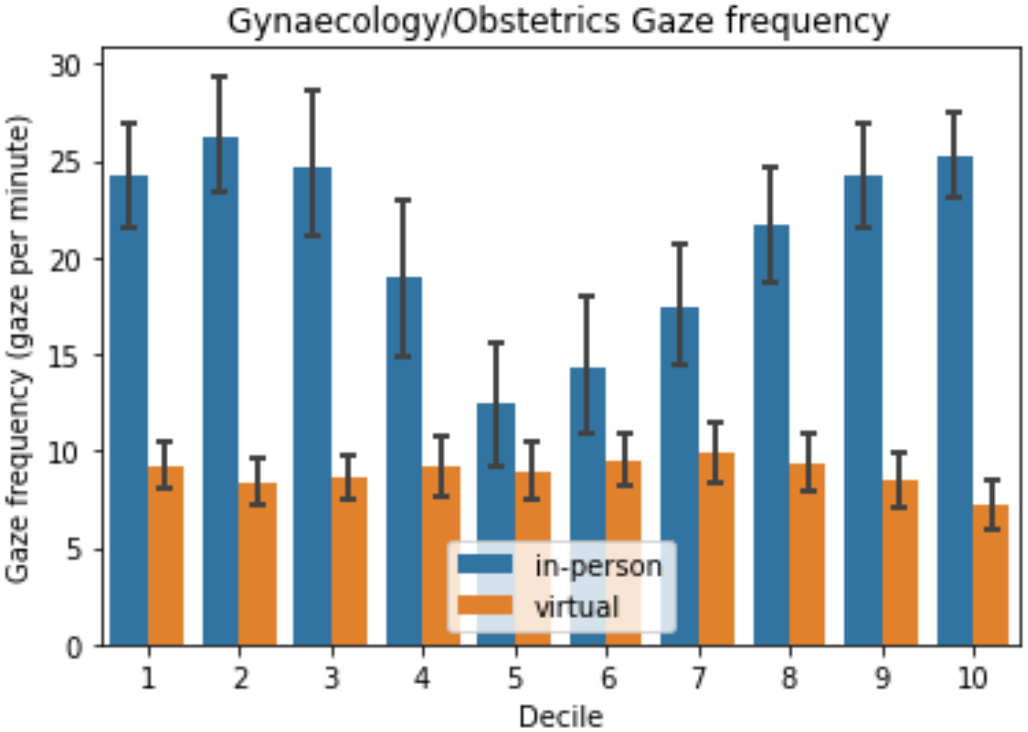
Gaze frequency across consultation time for Gynaecology/Obstetrics specialty. Consultation time is divided into 10 slots.

We also verify the dependence between gaze frequencies for both conditions. Gaze frequency of virtual consultations is dependent to in-person frequency (*P* = 0.0049). In this case the frequency is almost constant for the virtual consultations. Finally, in-person duration and frequency are highly correlated (*r* = 0.5707, *P <* 0.001), and virtual duration and frequency are not correlated (*r* = 0.185, *P <* 0.001).

#### 2) Endocrinology

Table II and Figure 8 show the total gaze time to the patient, for each doctor in this specialty. Note that D10 and D15 have significant differences, while D3 shows a tendency. The joint analysis of the three doctors shows a significant difference between in-person and virtual total gaze time (*P <* 0.001)

**TABLE II:** Endocrinology results and summary statistics.

| Doctor | *Med<sub>f</sub>* | *Med<sub>v</sub>* | *p-value* | Cohen's *d* |
|---|---|---|---|---|
| D3 | 37.7% | 40.8% | 0.11 | - |
| D10 | 27.7% | 42.2% | 0.01 | 0.40 |
| D15 | 44.6% | 58.2% | 1.6 × 10<sup>−5</sup> | 0.68 |
| Joint | 37.3%'s | 50.9% | 0.7 × 10<sup>−5</sup> | - |

**Fig. 8:**
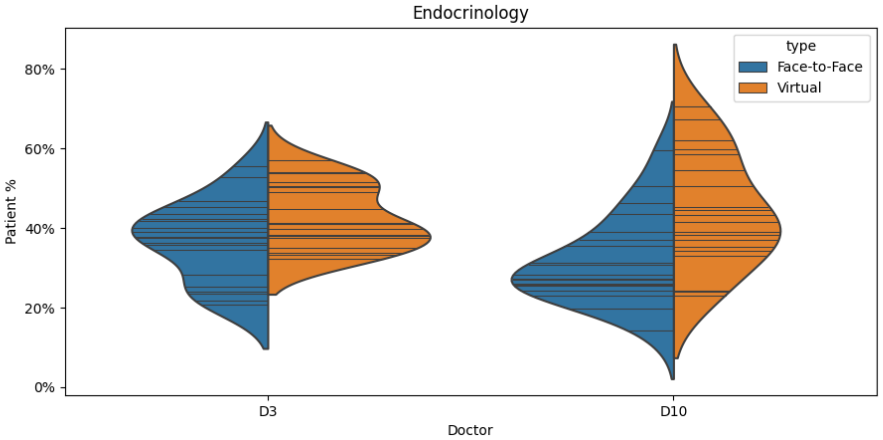
Endocrinology doctors violin plots Regarding gaze duration, Figure 9 shows the average duration of a fixation across all consultation deciles. Note that there is no trend of gaze duration for the virtual consultations (*β* = 0.155, *P* = 0.569). Similary, there is no clear trend for in-person consultations (*β* = 0.332, *P* = 0.141). We also verify the dependence between gaze duration for both conditions. Gaze duration of virtual consultations is independent to in-person frequency (*P* = 0.975).

**Fig. 9:**
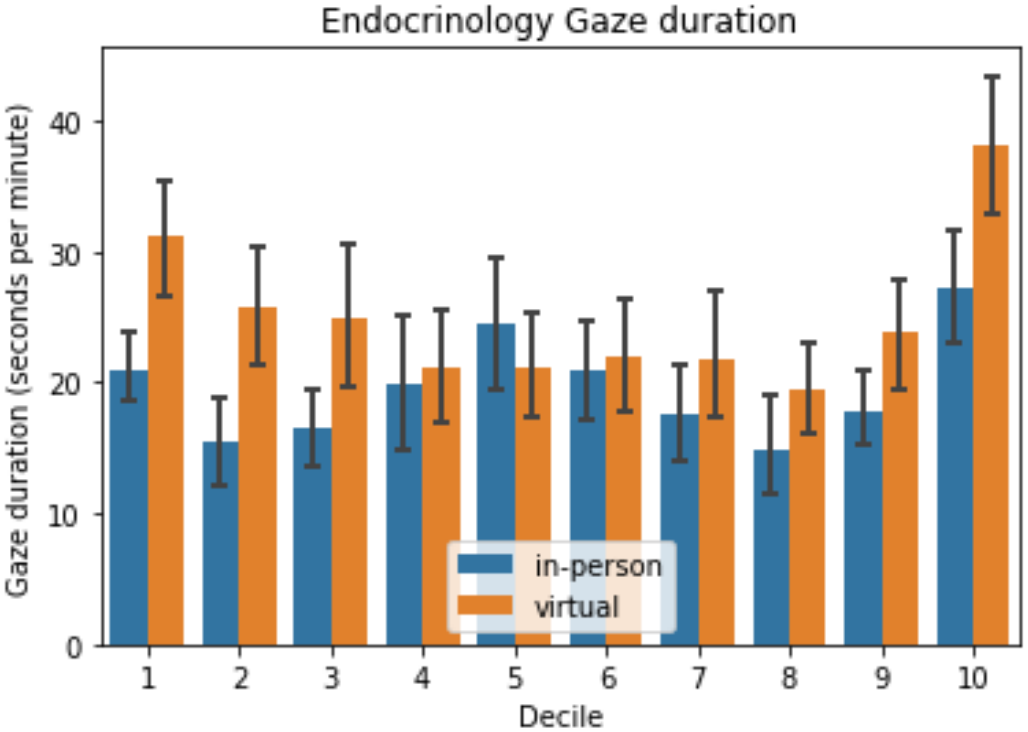
Gaze duration across consultation time for Endocrinology specialty. Consultation time is divided into 10 slots.

Regarding gaze frequency, Figure 10 shows the average gaze frequency across all consultation deciles. In the case of virtual consultations, there is no clear time-series trend (*β* = 0.0721, *P* = 0.439). Similary, there is no clear trend on in-person consultations (*β* = 0.0742, *P* = 0.401). In-person gaze duration decreases during the first half of the consultation, while increases during the second half of the consultation.

**Fig. 10:**
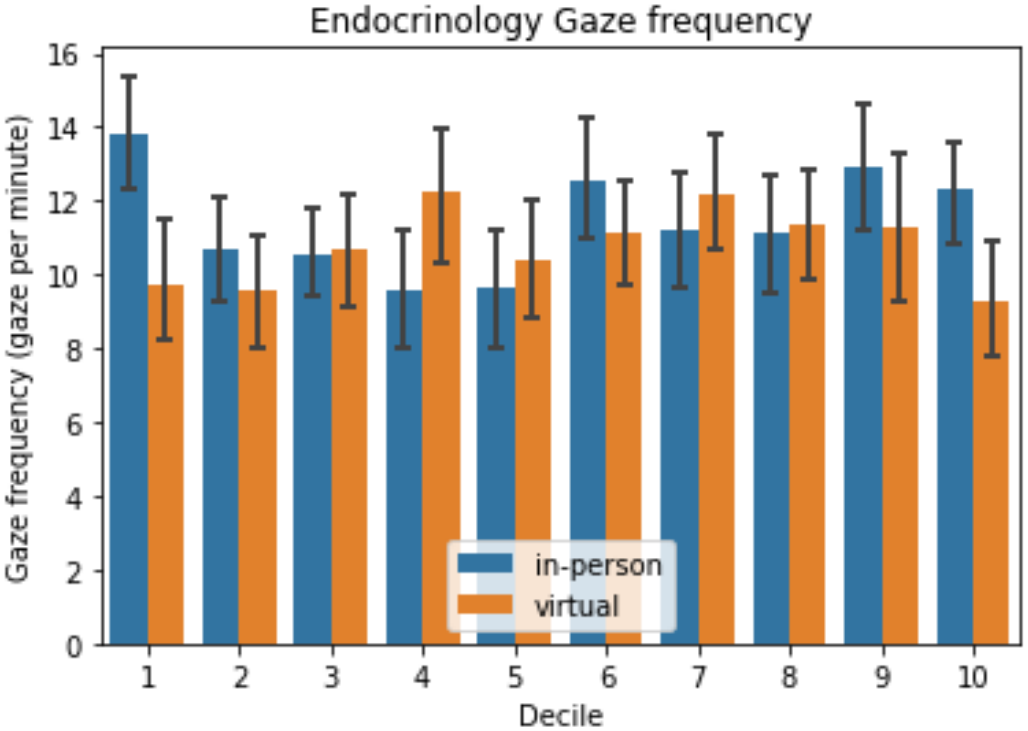
Gaze frequency across consultation time for Endocrinology specialty. Consultation time is divided into 10 slots.

We also verify the dependence between gaze frequencies for both conditions. Gaze frequency of virtual consultations is dependent to in-person frequency (*P* = 0.0049). In both cases, the frequency remains close to constant during the consultations. There is a significant but very small correlation for both types of consultations: (i) In-person duration and frequency are correlated (*r* = 0.27, *P <* 0.001), and (ii) virtual duration and frequency are not correlated (*r* = *−*0.073, *P* = 0.142).

#### 3) Neurology

Table III and Figure 11 show the total gaze time to the patient, for each doctor in this specialty. Note that D2 and D8 have significant differences, while D1 shows a tendency. The joint analysis of the three doctors shows a significant difference between in-person and virtual total gaze time (*P <* 0.001)

**Table III:** Neurology results and summary statistics.

| Doctor | *Med*<sub>*f*</sub> | *Med*<sub>*v*</sub> | *p-value* | Cohen's *d* |
|---|---|---|---|---|
| D1 | 45.8% | 48.3% | 0.18 | - |
| D2 | 54.9% | 65.9% | 3.1 × 10<sup>−3</sup> | 0.47 |
| D8 | 32.8% | 49.9% | 0.5 × 10<sup>−5</sup> | 0.72 |
| Joint | 44.7% | 57.7% | 2.1 × 10<sup>−5</sup> | - |

**Fig. 11:**
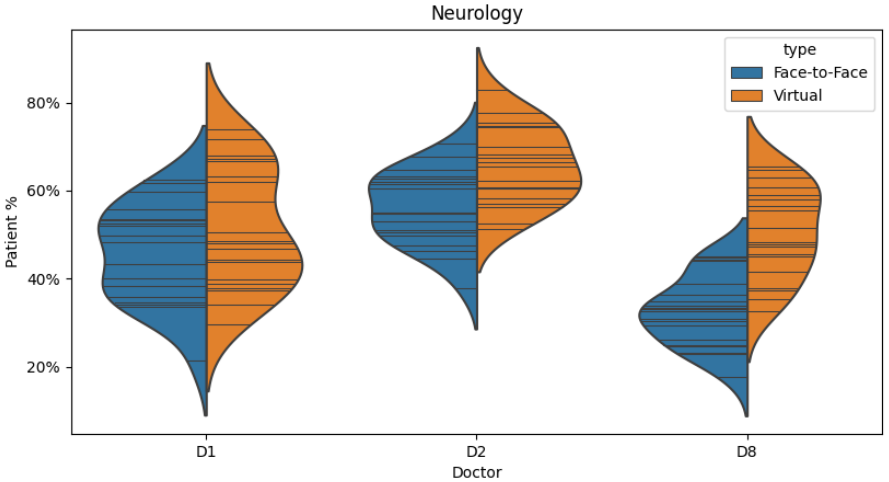
Neurology doctors violin plots

Regarding gaze duration, Figure 12 shows the average duration of a fixation across all consultation deciles. Note that there is no trend of gaze duration for the virtual consultations (*β* = *−*0.176, *P* = 0.407). Similarly, there is no clear trend for in-person consultations (*β* = *−*0.1079, *P* = 0.583). We also verify the dependence between gaze duration for both conditions. Gaze duration of virtual consultations is independent to in-person frequency (*P* = 0.96).

**Fig. 12:**
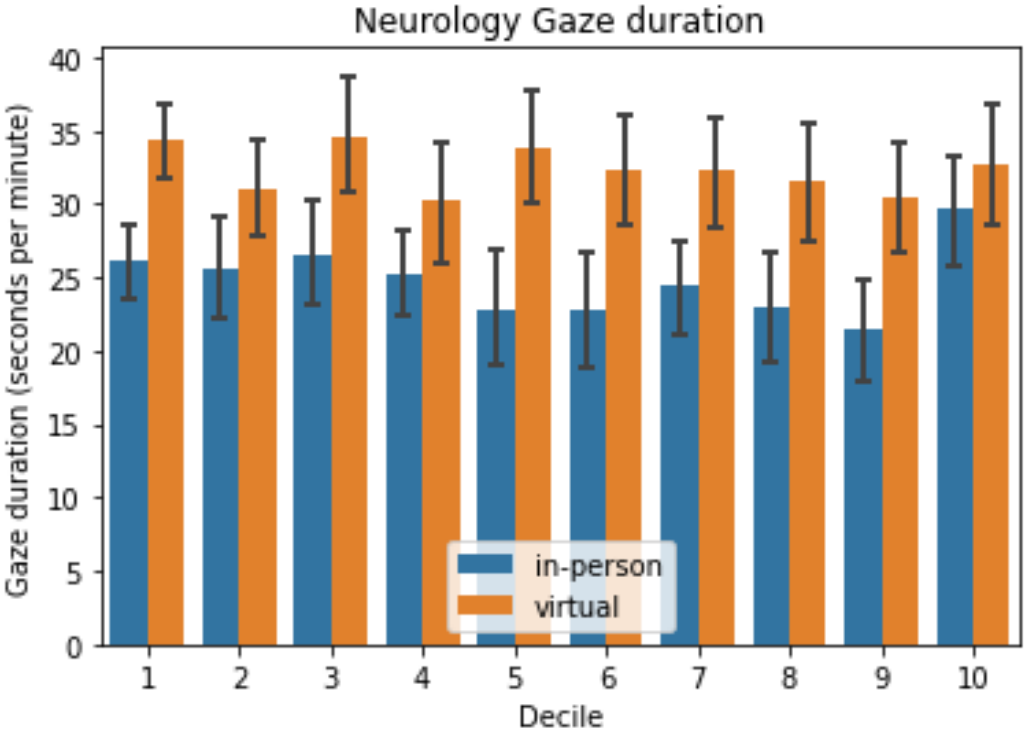
Gaze duration across consultation time for Neurology specialty. Consultation time is divided into 10 slots.

Regarding gaze frequency, Figure 13 shows the average gaze frequency across all consultation deciles. In the case of virtual consultations, there is a decreasing time-series trend (*β* = *−*0.1393, *P* = 0.026), while there is no clear trend on in-person consultations (*β* = 0.0019, *P* = 0.985).

**Fig. 13:**
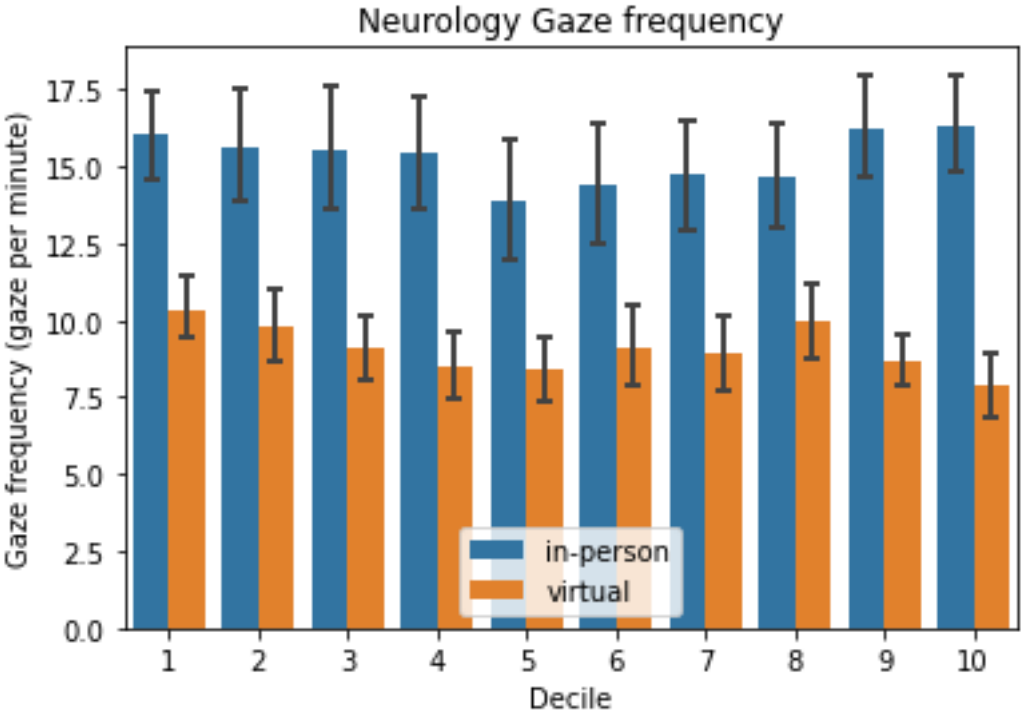
Gaze frequency across consultation time for Neurology specialty. Consultation time is divided into 10 slots.

We also verify the dependence between gaze frequencies for both conditions. Gaze frequency of virtual consultations is dependent to in-person frequency (*P* = 0.039). In both cases, the frequency remains close to constant during the consultations. There is a significant but small correlation for both types of consultations: (i) In-person duration and frequency are correlated (*r* = 0.248, *P <* 0.001), and (ii) virtual duration and frequency are not correlated (*r* = *−*0.039, *P* = 0.339).

#### 4) General and Family Medicine

Table 4 and Figure 14 show the total gaze time to the patient, for each doctor in this specialty. Note that D4 and D16 have significant differences, while D5 and D7 show a tendency. The joint analysis of the four doctors shows a significant difference between in-person and virtual total gaze time (*P <* 0.001).

**TABLE 4:** General and Family Medicine results and summary statistics.

| Doctor | *Med*<sub>*f*</sub> | *Med*<sub>*v*</sub> | *p-value* | Cohen's *d* |
|---|---|---|---|---|
| D4 | 38.2% | 43.2% | 0.049 | 0.31 |
| D5 | 62.4% | 59.3% | 0.19 | - |
| D7 | 46.4% | 55.7% | 0.11 | - |
| D16 | 43.6% | 59.2% | 0.0059 | 0.54 |
| Joint | 46.5% | 57.1% | 2.9 × 10<sup>−4</sup> | - |

**Fig. 14:**
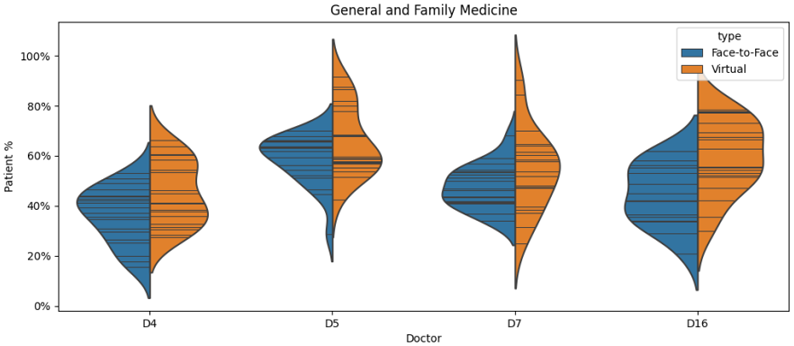
General and Family Medicine doctors violin plots

Regarding gaze duration, Figure 15 shows the average duration of a fixation across all consultation deciles. Note that there is a decreasing trend of gaze duration for the virtual consultations (*β* = *−*1.164, *P <* 0.001), while there is no clear trend for in-person consultations (*β* = *−*0.3005, *P* = 0.142). We also verify the dependence between gaze duration for both conditions. Gaze duration of virtual consultations is independent to in-person frequency (*P* = 1.0).

**Fig. 15:**
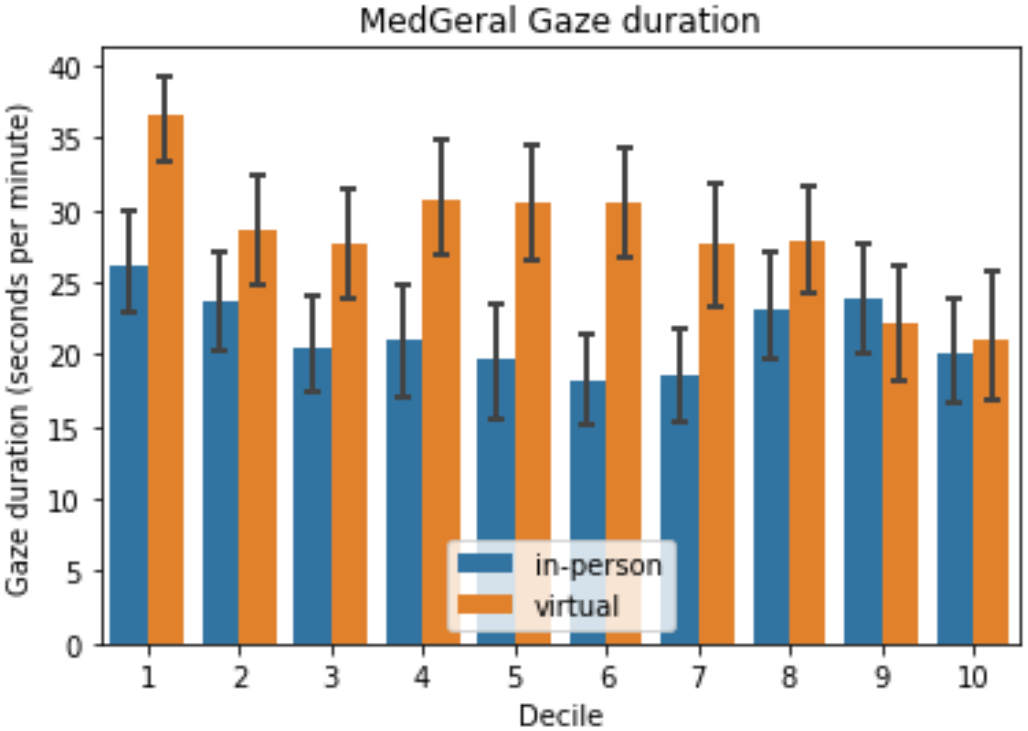
Gaze duration across consultation time for General and Family medicine specialty. Consultation time is divided into 10 slots.

Regarding gaze frequency, Figure 16 shows the average gaze frequency across all consultation deciles. In the case of virtual consultations, there is a decreasing time-series trend (*β* = *−*0.1203, *P* = 0.034), while there is no clear trend on in-person consultations (*β* = 0.0318, *P* = 0.694).

**Fig. 16:**
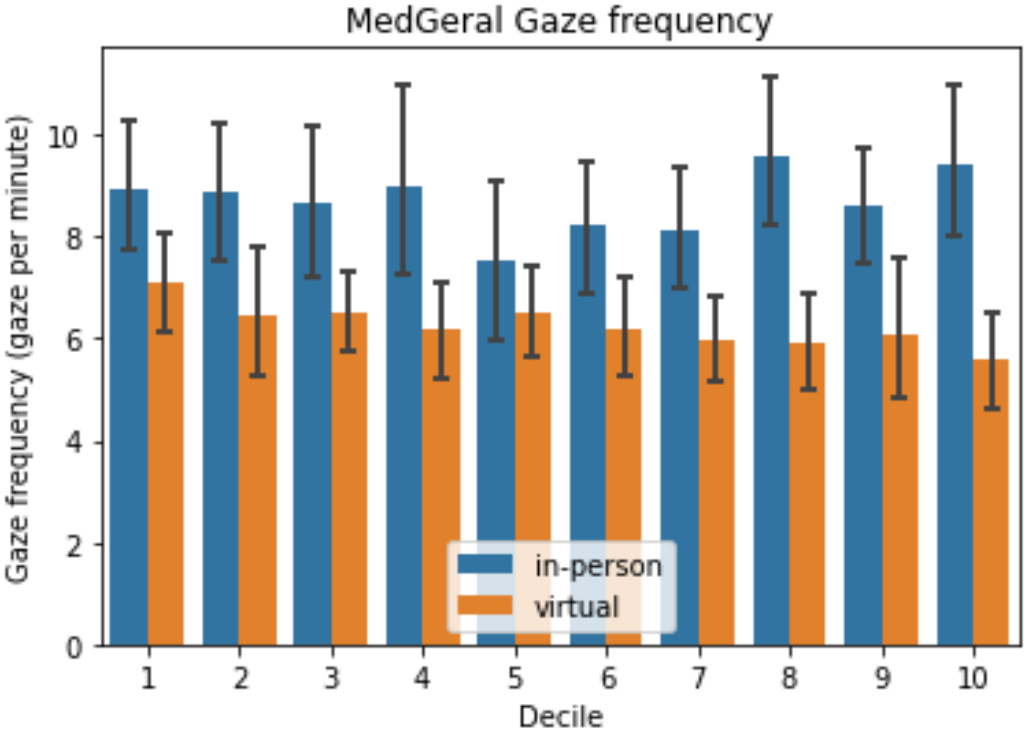
Gaze frequency across consultation time for General and Family medicine specialty. Consultation time is divided into 10 slots.

We also verify the dependence between gaze frequencies for both conditions. Gaze frequency of virtual consultations is independent to in-person frequency (*P* = 0.482). In both cases, the frequency remains close to constant during the consultations.

There is a significant but relatively small correlation for both types of consultations: (i) In-person duration and frequency are correlated (*r* = 0.357, *P <* 0.001), and (ii) virtual duration and frequency are correlated (*r* = 0.242, *P <* 0.001).

## VI. Discussion

Overall our findings provide new insights into the doctor-patient relationship in a digital era. More importantly, they provide a look at the effect of virtual consultations on the doctor-patient relationship, specifically on their effect on the doctor’s gaze behaviour. In three of the four medical specialties analyzed, we found a pronounced general tendency for doctors to look more at the patient during virtual consultations. However, the other medical specialty (Gynaecology/Obstetrics) seems to be the least affected by the consultation environment change. In this case, half of the doctors present no differences or tendencies between consultation environments, leading to the Gynaecology/Obstetrics specialty presenting very few tendencies between consultation environments. Regarding gaze duration and frequency, the global analysis found differences between the in-person consultations of our study and the ones presented in [26]. The decreasing pattern on both gaze duration and frequency observed in [26], was not found in our study. Other observation from [26], is that gaze frequency and gaze duration were not correlated. In our study both duration and frequency are correlated, for both types of consultations.

Concerning the individual doctors’ results, out of the fourteen participating doctors, seven showed statistically significant differences with medium-sized size effects, while another five showed tendencies to have higher *Patient%* in the virtual environment, with only two showing no differences in gaze behaviour. Overall, these results show the virtual consultation environment’s effect on the doctors from the study population. Additionally, these results also show the virtual consultation medium does not degrade eye contact since no doctor looked at the patient more during virtual consultations.

On the global analysis, we found that (i) gaze duration on virtual is independent from gaze duration in-person, (ii) gaze frequency on virtual is dependent on gaze frequency in-person. On the one hand, gaze duration has a different pattern between consultations and doctors adapt differently to the two environments. On the other hand, gaze frequency seem to have a very small variation between the two environments. We also found that for the same type of consultation, gaze duration has a very low correlation to gaze frequency. This trend also is verified in the per-specialty analysis.

On the per specialty analysis, we found that gaze duration on virtual is dependent to gaze duration in person for Gynaecology, Endocrinology. On the other hand, it is independent for Neurology and General Medicine. Gaze frequency on virtual consultations is dependent to frequency in-person for Gynaecology, Endocrinology and Neurology. General medicine is the only specialty where there is no gaze frequency dependence. In addition to assessing the virtual consultation environment’s effect on doctor gaze behaviour, this study also provides, to our knowledge, the first implementation of an appearance-based gaze estimation pipeline in the clinical setup using a low-cost webcam. This task came with many challenges, in addition to the usual robustness against head pose, glasses and environment changes, the gaze estimation system had to be able to deal with the obligatory use of masks. Since the use of masks was not common before the pandemic, all the gaze estimation datasets and systems were not tailored for the task of estimating the gaze of a mask-wearing subject. The solution, which consisted in using a different mask that did not occlude the mouth and nose region, worked remarkably well through the various tests performed and continued to work throughout the study. In the end, the gaze estimation pipeline provides an easy way to record and analyze the doctor’s gaze behaviour during consultations. It opens up many applications in the study of non-verbal communication in the doctor-patient relationship, some of which we will suggest in Section VII-A. The implemented methodology using an automated gaze estimation pipeline led to the capture of a much larger number of interactions than previous studies of gaze in applied settings. While [26] provided 100 clinical interactions divided between 14 doctors, our study provided 260 face-to-face clinical interactions and 260 virtual clinical interactions. The use of an automated gaze estimation pipeline also allowed us to study the gaze in more detail than with manually annotated gaze estimates. Additionally, using an appearance-based gaze estimation system like *Gaze360* provides a method that can be implemented in a variety of situations since it only needs an off-the-shelf webcam to be feasible. This clearly demostrates the scalability of the methodology.

Our study has some limitations. The first limitation relates to a caregiver being present and the patient in some face-to-face consultations recorded. The way we classify gaze estimates classified any gaze towards the caregiver as gaze towards the patient, which might lead to some overestimation. However, the goal of measuring the division of the doctor’s attention between screen and patient is still accomplished. The second limitation is related to the way we classify gaze direction, which leads to the *Patient%* metric being a estimation of the real time percentage the doctor spends looking at the patient. The zone used to define the *Patient* zone was the area on the side of the screen where the patient is located. Thus, the *Patient* zone not only defines where the patient is located but also the surrounding area. The assumption used to make this division is that whenever the doctor looks to the side of the screen where the patient is, the doctor is looking at the patient. This is a strong assumption which leads to very good estimations of the real time the doctor spends looking at the patient through the *Patient%* metric. This fact was explored in [31], where the authors explored that physicians can simulate eye contact by gazing at the camera. Doing so improves their communication and interpersonal skills ratings and could potentially enhance their communication with their patients. Another possible issue that was accounted for before the start of the study is related to the Hawthorne effect. The Hawthorne effect is a psychological effect that refers to the change in subjects’ behaviours when they know they are being observed. To mitigate these effects, we kept the actions the doctor had to make that did not follow the regular consultation routine to a minimum, which is accomplished by only asking the doctor to click the *Record/Stop* button in the Recording interface in the beginning and at the end of the consultation. The successful implementation of the gaze estimation pipeline allows us to analyze doctors’ gaze behaviour in more detail and robustly than before. This ability is crucial to analyzing the impact of non-verbal communication in the physician-patient relationship to improve the quality and efficiency of the health care provided. The applications of gaze estimation in the clinical setup are immense. It could be used in the assessment of clinical setup changes. We assessed the impact of virtual consultations against face-to-face consultations. However, many other setup changes can also be analyzed. Software updates can be analyzed to increase doctors’ efficiency when navigating the interfaces on the computer. Consultation room changes impact on doctor’s gaze behaviours like lighting conditions changes and patient positioning changes can be analyzed thoroughly. More interestingly, gaze estimation can be used for digital biomarker analysis. During a consultation, the patient’s attention can be analyzed to see how it evolves throughout the visit. Children’s gazes during pediatric therapy sessions could be analyzed to assess if they are focusing on what the therapist wants and improve the overall therapy sessions in the future. Like pediatric therapy, patients with neurological/psychological disorders gaze could be studied during therapy sessions to assess the effectiveness of the therapy sessions and possibly improve them in the future.

Even though these results are not enough to make robust generalizations about medical specialties as a whole, they still provide an insight into the effect of virtual consultations on doctor behaviour. Consequently, these promising initial results deserve further study and corroboration to better understand the doctor-patient relationship.

### VII. Conclusions

This study implemented a scalable and low-cost 3D appearance-based gaze estimation pipeline in the clinical setup. Additionally, this technological advancement enabled a thorough evaluation of how virtual consultations influence the duration for which doctors in the study group maintain eye contact with their patients. In addition, gaze duration during the virtual consultations is larger on the majority of the consultation deciles. Gaze frequency did not show significant differences between virtual and in-person consultations. Doctors spend significantly more time looking at the patient during virtual consultations, with a few exceptions. Three out of the four specialties present prominent tendencies to look more at the patient during virtual consultations when compared to face-to-face consultations and none of the specialties looked less to the patients, suggesting that virtual consultations did not negativelly impact the eye contact between doctor and patient.

### A. Future Work

The impact of virtual consultations on doctors’ gaze behaviour is apparent. However, it should continue to be analyzed to provide us with a better understanding of the consequences to the physician-patient relationship. Therefore, a large scale study involving more doctors with a higher number of consultations recorded should be performed. Such a study is essential to support and corroborate these initial results obtained to fully understand the impact of virtual consultations on doctors’ gaze behaviour. In addition to a larger amount of samples collected, this future project could also include the extraction of more gaze features along with the *PoG*s, in the form of saccades, smooth pursuits or even face landmarks. This would allow for a deeper study of the doctor’s gaze patterns.

Another work that could be done in support of this study is on the correlation between the *Patient%* metric and the actual percentage of time the doctor spent looking at the patient. To obtain the actual percentage, the doctor has to put on the eye-trackers that look like glasses, which provide images of the Field-Of-View (FOV) of the doctor and the gaze located in the FOV image (method used in [26]). Finding the exact relationship between the *Patient%* and the actual percentage of time is an important step to support of our results.

Additionally, we need to study more than gaze to evaluate the total impact of non-verbal communication in the physician-patient relationship. Therefore, coupling gaze estimation with other types of machine learning algorithms to analyze other non-verbal cues like voice pitch, monotony, facial expressions, or body posture is the next step in the studying the impacts of non-verbal communication in the physician-patient relationship.

## Data Availability

The data is not available

